# Effect of Respiratory Muscle Training on Dysphagia in Stroke Patients - A Retrospective Pilot Study

**DOI:** 10.1101/2020.02.08.20021303

**Authors:** Robert J. Arnold, Nina Bausek

## Abstract

**Background:** Dysphagia is prevalent with cerebrovascular accidents and contributes to the burden of disease and mortality. Strengthening of the dysfunctional swallow muscles through respiratory muscle training (RMT) has proven effective in improving swallow effectiveness and safety. However, approaches to strengthen only the expiratory muscle groups (EMST) dominate the clinical study literature, with variable outcomes. This study investigated the effect of a simultaneous inspiratory and expiratory muscle strengthening strategy to improve swallowing function in stroke patients.

**Methods:** Twenty post-stroke patients were randomly assigned to either intervention group (IG) or control group (CG). The intervention group was treated with three 5-minute sessions of resistive respiratory muscle training every day for 28 days, while the control group received no RMT. Respiratory and swallow outcomes were assessed pre- and post-intervention and included Mann Assessment of Swallowing Ability (MASA), Fiberoptic Endoscopic Evaluation of Swallowing (FEES) with Penetration/Aspiration Scale (PAS), Functional Oral Intake Scale (FOIS), patient visual analogue scale (VAS), and peak expiratory flow (PEF).

**Results:** After 28 days, the intervention group demonstrated greater improvements (pVal < 0.05) in PEF (IG: 168.03% vs CG: 17.47%), VAS (IG: 103.85% vs CG: 27.54%), MASA (IG: 37.28% vs CG: 6.92%), PAS (IG: 69.84% vs CG: 12.12%), and FOIS (IG: 93.75% vs CG: 21.21%).

**Conclusion:** Combined resistive inspiratory and expiratory muscle training is a feasible and effective method to improve signs and symptoms of dysphagia in stroke patients.

## Introduction

The physiological process of deglutition comprises a complex set of events which begin when a given food, liquid, and/or medication bolus is introduced into the oral cavity via the lips and ends once the same bolus empties from the esophagus via the lower esophageal sphincter into the stomach [1].

The signs and symptoms of swallow pathophysiology may include but are not limited to prolonged mastication, food/liquid/medication residuals in the oral cavity after the swallow, impaired labial containment of secretions as well as portions of a given food, liquid, and/or medication bolus, nasal regurgitation, a delay in the onset of the swallow, coughing, choking, regurgitation of bolus materials after a swallow, globus, and weight loss [2]. For the purpose of this paper, dysphagia is defined as dysfunctional swallow physiology involving the oral cavity, velopharyngeal port, larynx, pharynx, esophagus, and/or the lower esophageal sphincter. This is consistent with the definition provided by the American Speech-Language-Hearing Association (ASHA) [3].

Although dysphagia can occur across the lifespan from neonates to geriatrics, it is more prevalent in the elderly (over 65) due to age-mediated muscle degeneration, and in those with neurological disorders such as stroke, multiple sclerosis (MS) and Parkinson’s disease (PD) [4,5]. While dysphagia affects 11% of elderly Americans living in communities, prevalence rises to 25% in hospitalized individuals and to 55% in people living in elderly care facilities. Other sources report an average prevalence of 13.5% to 22.6% in unselected adults, almost half of which have not discussed their recurrent swallow problems with a physician. In patients with underlying neurological disorders, the prevalence is significantly higher up to 64% in stroke patients, 34% in MS and 81% in PD patients [4,6,7].

Consequences of dysphagia include malnutrition, dehydration, and weight loss, as well as increased risk of cardiac and respiratory conditions. Dysphagia is associated with a 13% increase in mortality, and causes 60,000 deaths per year in the US due to complications, especially aspiration pneumonia. Following a stroke, aspiration pneumonia increases the risk of mortality as well as the average cost of hospitalization by $27,633 [8].

Furthermore, dysphagia poses a significant burden on the healthcare system due to prolonged hospital stays, and the need for passive enteral feedings in affected patients. In the US, the current annual economic burden is estimated at $547 million for prolonged hospitalization and $670 million for enteral feeding supplies, amounting to total annual costs well in excess of $1 billion, with an expected annual increase due to the aging population [4,9]. In addition, patients over 65 presenting with dysphagia, choking or globus represent 37.6% of all emergency department visits for food-mediated adverse events [10].

Treatment of dysphagia reflects the complexity of the disorder, and requires a multidisciplinary approach. Early intervention and therapy is associated with reduced risk of aspiration pneumonia and quicker recovery [11,12].

Both the muscles of inhalation as well as the muscles of exhalation are critical integrated components of the upper aerodigestive tract, whose function is necessary for swallow physiology [13]. Strengthening of either muscle group by inspiratory muscle strength training (IMST) or expiratory muscle strength training (EMST), respectively, has shown positive effects on respiration as well as on swallow physiology [14–17]. Some studies have also shown an exercise regimen which includes both IMST and EMST as separate components of the treatment plan to be beneficial in maximizing airway safety with swallowing in neurological patient populations [18,19]. The investigators elected to utilize a combined resistive IMST and EMST device (The Breather™, PN Medical Inc., US Patent Number 4,739,987)[20], thus allowing for simultaneous strengthening of the muscles of inhalation and exhalation [21,22]. This retrospective pilot study analyzes the effectiveness of a 4 week combined respiratory muscle training (cRMT) program on swallow function in stroke patients with diagnosed dysphagia following a single cerebrovascular accident (CVA). This research received an IRB exemption - HIPAA full waiver of authorization and regulatory opinion from the Western Institutional Review Board.

## Methods

Data analyzed in this study is derived from persons who resided in the State of Alabama whose health had been compromised by a single acute CVA and who lacked a pay source to cover the cost of swallow therapy. Each subject had been referred to *pro bono* clinical Speech-Language Pathology (SLP) services as needed through the Office of Hispanic Ministry at a Catholic Church in the Greater Birmingham Area of Alabama. Data was collected between 1998 and 2009.

Subjects who met the inclusion criteria presenting with dysphagia after a cerebrovascular accident (CVA) were included in this study. Inclusion criteria comprised of no prior neurological history, no history of swallow deficits prior to CVA, no pharmacological or surgical interventions, and no other therapy for swallowing during the study period. Exclusion criteria were failure to meet one or more of the inclusion criteria. In addition, there was no therapy of any other kind, including physiotherapy, occupational therapy, respiratory therapy, or speech therapy. Subjects were controlled for age, sex, race, hemisphere of CVA, presence of single CVA without extension. While all patients remained on the referral waiting list for treatment, each patient was offered a choice to be trained in the use of The Breather™. Subjects who chose to use cRMT were considered as the intervention group (IG), while patients who chose to not use cRMT were treated as the control group (CG) for analysis.

### cRMT Intervention

Combined respiratory muscle training (cRMT) was performed using a resistive inspiratory and expiratory muscle training device. Patients were instructed to wear a nose clip, sit upright, and to forcefully inhale and exhale through the device using diaphragmatic breathing technique. Patients unable to maintain a tight lip seal with the mouthpiece of the Breather were trained to utilize a CPR (cardiopulmonary resuscitation) facemask in lieu of the standard Breather mouthpiece. The duration of intervention was 28 days (4 weeks), and included one skilled intervention and 6 homework sessions per week. During the skilled intervention, patients performed 3 sets of 1 to 5 minutes of cRMT under therapist supervision. The home-based sessions were performed without supervision and consisted of 3 sets of a maximum of 5 minutes of cRMT per session, with 3 sessions per day. cRMT intensity was defined as the highest tolerated settings for both inhalation and exhalation. Importantly, these settings may be set independently from each other. Compliance and training adherence was monitored via patient communication during weekly skilled intervention sessions. The control group only received recommendations pertaining to the safest food consistencies, liquid consistencies, and medication consistencies, as well as positioning and compensatory strategies to optimize airway safety with swallowing while awaiting standard of care treatment.

### Assessments

Baseline and final assessments were identical. These included Mann Assessment of Swallowing Ability (MASA)[23], Fiberoptic Endoscopic Evaluation of Swallowing (FEES) [24] with Penetration/Aspiration Scale (PAS)[25], Functional Oral Intake Scale (FOIS)[26], patient’s visual analogue scale (VAS)[27], and peak expiratory flow (Spir-O-Flow Peak Flow Pocket Monitor, Spirometrics).

### Data Analysis

Data was analyzed using SPSS and StatPlus. A two-tailed t-test was selected for analysis as the direction of the difference could not be predicted on the basis of available relevant research.

## Results

### Patient history and demographics

In total, data from 20 subjects who either had or had not used the Breather fulfilled the inclusion criteria and were used for analysis in the study. Subject data were allocated to either intervention or control group. Based on the medical history and physicals reviewed for each subject, 9 subjects had left hemisphere infarction, 7 subjects had right hemisphere infarction, and 4 subjects had sustained brainstem infarction.

Demographics and baseline assessment data are outlined in table 1.

**Table 1:** Patient demographics

|  | Intervention Group | Control Group |
| --- | --- | --- |
| **n** | 10 | 10 |
| **Sex male** | 2 | 6 |
| **Sex female** | 8 | 4 |
| **Mean age** | 70.50 | 66.10 |

The baseline scores for all variables were compared in terms of experimental condition, and gender, using MANOVA. The multivariate test was not significant for experimental group, F(5,9)=2.15; p=.151, or for gender, F(5,9)=1.12, p=.415. Therefore, no covariates were included in the main analysis.

Baseline data were collected before the initiation of the cRMT intervention and is outlined in table 2.

**Table 2:** Data collected before cRMT intervention at baseline. Abbreviations: p: p-Value (T-Test, two tailed), PEF: peak expiratory flow, VAS: visual analog scale, MASA: Mann Assessment of Swallowing Ability, PAS: Penetration Aspiration Scale, FOIS: Functional Oral Intake Scale.

| Baseline data: | Intervention Group | Control Group | p between groups |
| --- | --- | --- | --- |
| PEF | 94 | 88 | 0.73142 |
| VAS | 44.10 | 37.40 | 0.56885 |
| MASA | 133.30 | 127.10 | 0.60229 |
| PAS | 6.30 | 6.60 | 0.70254 |
| FOIS | 3.20 | 3.30 | 0.91152 |

Analysis of baseline data shows that there are no significant differences between the intervention group and the control group (p value between groups).

Posttest final assessment was performed after 4 weeks of cRMT intervention, and is summarized in table 3.

**Table 3:** Data collected at the end of the 4 week intervention. Abbreviations: IG: intervention group, CG: control group, SD: standard deviation, p: p-Value (T-Test, two tailed), PEF: peak expiratory flow, VAS: visual analog scale, MASA: Mann Assessment of Swallowing Ability, PAS: Penetration Aspiration Scale, FOIS: Functional Oral Intake Scale.

|  |  | Baseline |  | Final |  |  |  |  |
| --- | --- | --- | --- | --- | --- | --- | --- | --- |
| Assessment | Group | Mean | SD | Mean | SD | Percent Change | p pre<post | p between groups |
| PEF | IG | 94 | 44.02 | 258.00 | 54.12 | 168.03% | <0.001 | <0.001*** |
|  | CG | 88 | 41.85 | 107 | 31.29 | 17.47% | .02240 |  |
| VAS | IG | 44.10 | 24.84 | 89.90 | 12.01 | 103.85% | <0.001 | .00127** |
|  | CG | 37.40 | 27.60 | 47.70 | 23.35 | 27.54% | .04355 |  |
| MASA | IG | 133.3 | 23.35 | 183.00 | 11.09 | 37.28% | <0.001 | <0.001*** |
|  | CG | 127.1 | 25.90 | 135.90 | 21.75 | 6.92% | .00762 |  |
| PAS | IG | 6.30 | 2.21 | 1.90 | 1.37 | 69.84% | <0.001 | .00158** |
|  | CG | 6.60 | 1.90 | 5.80 | 1.87 | 12.12% | .08684 |  |
| FOIS | IG | 3.20 | 2.20 | 6.20 | 0.92 | 93.75% | <0.001 | .01153** |
|  | CG | 3.30 | 2.16 | 4.00 | 1.89 | 21.21% | .11076 |  |

Average peak flow increased significantly by 168.03% in the intervention group (IG), compared to 17.47% in the control group (CG). The visual analog scale (VAS) improved by 103.85% in the intervention group, and 27.54% in the control group. Likewise, scores in the assessment of swallowing ability (MASA) also significantly improved by 37.28% in the intervention group, compared to a 6.92% increase in the control group. The penetration/aspiration scale (PAS) was reduced by 69.84% in the intervention group, and by 12.12% in the control group. A reduction in PAS numerical value signifies an improvement in swallowing safety by indicating a reduction in airway invasion. The food oral intake scale (FOIS) improved by 93.75% in the intervention group, and by 21.21% in the control group, indicating improvement in the functional level of intake of food and liquid. Comparison of baseline and final evaluation data within the groups revealed significant improvements over time in all assessments in the intervention, but not the control group. Between group analysis showed either two-star (**, pVal<0.05) or three-star (***, pVal<0.001) significance in all evaluated parameters, indicating the effectiveness of the intervention in improving swallowing-related symptoms.

## Discussion

The data analysis presented reveals the benefits of the utilization of cRMT as an effective therapy in the treatment of neurogenic dysphagia following CVA. Four weeks of using a combined resistive RMT therapy device significantly improved airway safety with swallow function, compared to no direct treatment. Clinical parameters that improved included both objective measures such as PEF, FEES and FOIS, as well as subjective evaluations such as the VAS and MASA. These findings underline the effectiveness of the applied RMT method in the setting of dysphagia following stroke.

Subjects evaluated in this study had employed a resistive combined inspiratory and expiratory muscle trainer that provides training resistance by breathing through different sized apertures covered by a silicone membrane. This method differs from the also widely distributed threshold devices. In contrast to previously reported different outcomes related to the type of device, recent research has revealed no significant differences in treatment outcomes when controlling for threshold dependent RMT devices and non-threshold dependent RMT devices. Some of the evidence highlighted increased cost-effectiveness and simpler instructions for usage of non-threshold RMT devices compared to threshold dependent RMT devices [28–30]. As the application of cRMT is likely to be more comprehensive than using IMST and/or EMST separately or sequentially, it may therefore be less cumbersome and more cost effective to use cRMT in lieu of IMST and/or EMST alone.

Respiratory muscle weakness (RMW) and associated swallow dysfunction is prevalent in stroke patients, with a reduction in respiratory muscle strength by approximately 50%, compared to matched controls. These deficiencies may contribute to tracheobronchitis and pneumonia due to aspiration, decreased cough effectiveness and poor pulmonary hygiene. On average, post-stroke pneumonia affects 10% of all patients, on average, and is associated with higher mortality, hospitalization rates, worse outcomes, and higher care needs [31,32].

Respiratory muscle training is an evidence-based strategy targeting RMW. It follows the principle that respiratory muscles respond to exercise and training stimuli in the same manner as skeletal muscles do, a notion that was originally observed by the Greek physician Claudius Galen [33], and that has inspired the development of systematic training of the respiratory muscles [34]. Regular workload by training against resistance triggers respiratory muscle hypertrophy, fiber formation, and improved function. This improves the adaptability of respiratory muscles to increased ventilatory demand, and/or the functionality of laryngeal and pharyngeal muscles used for speech and swallow.

This study showed that respiratory muscle strengthening of both the inspiratory and expiratory muscles significantly improves peak flow and swallowing function, as assessed by swallowing ability, food intake and penetration/aspiration, in patients with CVA. 4 weeks of cRMT improved peak flow by 168%, penetration/aspiration by 70%, food oral intake by 94%, and swallowing ability by 37% (table 2).

Improvements in peak expiratory flow may correlate with more efficient clearance of supraglottic and tracheal secretions or aspirated material, which is also reflected by improved PAS scores. Associated improvements in deglutition can be expected to reduce pneumonia and malnutrition. Furthermore, improved peak expiratory flow may contribute to reduced risk of mortality, as peak flow has recently been identified as an independent predictor of mortality in the aging population [35].

The overall findings are in agreement with Pitts et al (2009), in that RMT of the expiratory muscles improved swallowing and cough function in patients with PD, including cough acceleration for improved airway clearance and increased penetration/aspiration scores, indicating improved efficacy during swallowing. A 4 week intervention of EMST improved the penetration/aspiration scores by 41%, and cough acceleration volume by 103% [36]. In contrast, other studies have not found EMST to be beneficial in post-stroke patients [32].Differences in outcomes may be attributed to different training regimes and adherence during the study period, and may also be device dependent. Furthermore, unilateral exercise limited to strengthening of the expiratory muscle group may limit effectiveness of this approach as an adjunctive approach for neurogenic swallow physiology rehabilitation. Further research is warranted.

### Limitations

There are some limitations inherently involved in this study. First amongst the limitations is the lack of randomization and the inability to have a placebo control group. This first limitation is associated with an inability to rule out skewing of the data towards positive results as motivation may have been higher in the intervention group.In addition, as the level of cognitive communication competence was not controlled, it may be the persons in the intervention group had a greater capability to process, attend, and implement the cRMT protocol. Furthermore, it is possible that persons who chose to participate in the intervention group were more active physically and had a higher level of cognitive communication competence than those who chose not to, thereby causing a difference in levels of exercise performed between the groups. As general exercise alone can improve symptoms of respiratory muscle weakness, the positive outcomes of this study may have been enhanced by this imbalance. Next, the positive results of the study were derived from a small population, possibly limiting its general conclusiveness. Therefore, conducting a double-blinded randomized prospective sham controlled trial with a larger sample population is recommended to confirm the results of this analysis. Future investigations evaluating the effect of cRMT on dysphagia following a single CVA may benefit from diversification of biophysical measurements by including the Yale Pharyngeal Residue Severity Rating Scale, as well as manometric measurements of respiratory muscle strength (maximum inspiratory and expiratory pressure, respectively) [37].

## Conclusion

This study analyzes the use of a resistive device which trains both inspiratory as well as expiratory muscle strength, while comparable studies may have employed devices that add workload to only one part of the breath cycle, or that provide insufficient workloads to elicit respiratory muscle hypertrophy. These approaches may be insufficient, as evidence reveals that functionality of both aspects of the breath cycle is required for effective cough and pulmonary hygiene [38]. This study therefore confirms feasibility, usefulness, and effectiveness of combined inspiratory and expiratory muscle training (cRMT) in people with dysphagia. Further investigations evaluating the effect of cRMT on dysphagia are warranted to verify the findings of the studies in larger and more diverse populations.

## Data Availability

As this study analysed historic data that was collected in a non-digital format in a pro bono clinic treating the uninsured, only manual data is available. Scanned data subsets can be made available upon request.

## Acknowledgments

The authors thank Sigfredo Aldarondo, MD, Tom Berlin, DHSc, MSc, RRT, and Roxann Diez Gross, PhD, CCC-SLP, for helpful comments and critical reading of the manuscript.

## Disclosures and Contributions

NB serves as independent Chief Scientist for PN Medical. RJA declares no conflict of interest.

## Notes

### Competing Interest Statement

Nina Bausek serves as independent Chief Scientist for PN Medical. Robert J Arnold declares no conflict of interest.

### Clinical Trial

This retrospective study was conducted under an IRB waiver granted from Western IRB.

### Funding Statement

This study did not receive any external funding.

